# Plasma Proteomic Signature of Mucolipidosis Type IV

**DOI:** 10.1101/2024.07.29.24311030

**Authors:** Brendan Tobin, Albert Misko, Victoria Miller-Browne, Madison Sangster, Yulia Grishchuk, Levi B. Wood

## Abstract

Mucolipidosis IV (MLIV) is an autosomal-recessive pediatric disease that leads to motor and cognitive deficits and loss of vision. It is caused by the loss of function of the lysosomal channel transient receptor potential mucolipin-1, TRPML1, and is associated with an early brain phenotype consisting of glial reactivity, hypomyelination, lysosomal abnormalities, and increased cytokine expression. Although the field is approaching the first translationally relevant therapy, we currently lack a molecular signature of disease that can be used to detect therapeutic efficacy. In the current study, we analyzed 7,322 proteins in the plasma proteome and compare protein profiles with clinical measures of disease severity (motor function, muscle tone, and age). To do so, we used aptamer-based protein profiling on plasma isolated from 18 MLIV patients and 37 aged-matched controls from a biorepository. We identified a total of 1,961 differentially expressed proteins between MLIV and control subjects, with functions spanning many major hallmarks of MLIV. Our analysis revealed a decrease in the abundance of neuronal proteins and an increase in muscle proteins, consistent with the neuronal dysfunction and muscle pathology observed in patients. In particular, lower levels of synaptic proteins (e.g., GABARAP) best correlated with disease severity. Next, we compared the plasma proteome of patients to the brain proteome from the mouse model of MLIV and identified shared alterations in 45 proteins. The up-regulated overlapping proteins were largely related to lysosomal function (e.g., ACTN2, GLB1), while the down-regulated proteins were largely related to myelination (e.g. TPPP3, CNTN2). Both signatures are consistent with our understanding of key disease hallmarks: impaired myelination and modified lysosomal function. Collectively, these data indicate that peripheral blood plasma protein signatures mirror changes found in the MLIV brain and suggest candidate markers relevant to MLIV pathology to be validated in future studies.

## Introduction

Mucolipidosis type IV (MLIV) is a neurodevelopmental and neurodegenerative disease that results from loss of function mutations in the lysosomal transient potential channel mucolipin 1 (TRPML1). Typically, patients exhibit delayed developmental milestones within the first year of life and then plateau in psychomotor function, consistent with that of an 18 to 20-month-old healthy child (1). As they progress through life, MLIV patients experience increased muscular hypertonicity, and deteriorating motor function (2, 3). Patients additionally develop a continuous decline in vision due to retinal degeneration and corneal clouding, ultimately leading to blindness (1, 3). One of the major hallmarks of the disease outside of the nervous system is high blood levels of gastrin that, together with developmental delay and corneal clouding, have been utilized as diagnostic criteria for MLIV. As clinically viable therapies for MLIV are being developed, it is critical to identify biomarkers capable of measuring therapeutic engagement in the central nervous system, which the diagnostic criteria cannot specifically measure. Thus, there is an essential need to broadly illuminate the effects of MLIV on blood proteomic profiles related to the primary disease pathology in the brain.

Brain pathology in MLIV is consistent with a hypomyelinating leukodystrophy (4). Notably, these pathologies are accompanied by progressive iron accumulation in the basal ganglia, cerebellar atrophy(5), accumulation of electron-dense lysosomal inclusions in all cell types (6) and reactive astrocytes and microglia (7, 8). Moreover, brain imaging of MLIV patients demonstrates a shortage of subcortical white matter, hypoplasia and dysgenesis in the corpus callosum, and variable white matter lesions (5).. Over time, patients experience a decrease in subcortical white matter and cerebellar volumes, while cortical gray matter volumes remain relatively intact (9). A few MLIV patients exhibit milder neurological impairment, which is associated with residual TRPML1 function demonstrated by certain allelic variants (2, 10).

MLIV is inherited in an autosomal recessive manner and caused by pathological variants in *MCOLN1*, the gene encoding TRPML1 (11–13). TRPML1 regulates multiple lysosomal functions and processes, including Ca^2+^-dependent lysosomal membrane fusion events in endocytosis, exocytosis, and autophagy (14); regulation of the transcription factor EB and the CLEAR gene network (15), autophagosome membrane assembly (16), and chaperon-mediated autophagy (17).

Loss of *Mcoln1* expression in mice mimics human MLIV and results in motor and cognitive deficits, accompanied by the characteristic brain pathology hallmarks such as hypoplastic corpus callosum, hypomyelination, glia activation and lysosomal storage inclusions (18–21). Motor deficits in mice become evident at 2 months of age and progress to hind-limb paralysis and premature death at around 7-8 months of age (21, 22). Histological assessment of the end-stage brain pathology showed partial loss of cerebellar Purkinje cells, with otherwise largely preserved neuronal populations (19, 20).

At present, clinical intervention for MLIV is limited to palliative care and management of some of the symptoms; however, CNS-targeted AAV-mediated gene transfer of *MCOLN1* was recently shown to restore or prevent motor decline in *Mcoln1^-/-^* mice and ameliorate brain pathology(22). These preclinical data provide a new hope for developing disease-altering therapies for MLIV. To help advance translational studies of MLIV and future clinical trials, we set out to define a blood plasma proteomic profile of MLIV as a first step toward molecular biomarker discovery.

In this study we identify a proteomic profile in plasma of MLIV patients and age- and sex-matched healthy controls using a multiplex protein quantification technology SomaScan. Using DNA-based Somamers that are specific to each protein target, the SomaScan assay delivers readouts of more than 7000 high and low abundance proteins from a single small quantity serum or plasma sample. We use these data to establish a protein signature of human MLIV, perform pathway analysis, and correlate differentially expressed proteins and pathways with motor function scores in patients to identify protein signatures related to neuro-motor disability and disease progression. We then compare human plasma and MLIV mouse brain proteomes and identify common overlapping protein signatures, providing further insights into disease mechanisms, and identifying protein biomarker candidates for future validation.

## Methods

### Study Design and Population

Patients with MLIV (N=17, F=9, M=8) were recruited through the Mucolipidosis Type 4 Foundation. Written informed consent was obtained from legal guardians for participation in our approved natural history and biomarkers studies according to protocols approved by the Massachusetts General Hospital Institutional Review Board. Written informed consent or assent was obtained from patients if their neurological capacity allowed. Inclusion criteria for patients included a documented diagnosis of MLIV by 1) clinical or research-based sequencing of *MCOLN1* and identification of two pathological *MCOLN1* alleles or 2) presence of the expected constellation of clinical symptoms associated with MLIV and documentation of at least one of the following: one pathological *MCOLN1* allele, elevated gastrin levels, or a tissue biopsy with evidence of lysosomal inclusions consistent with MLIV. MLIV cases were assigned to either typical (N=15) or mild (N=2) presentation. Based on our clinical experience, we classified patients as mild if they had obtained independent ambulation at some point in life. All patients underwent a full examination by a single board-certified pediatric neurologist, and their function scored with the Brief Assessment of Motor Function (BAMF) scales (gross motor, upper extremity gross motor, fine motor, deglutition, and articulation) and Modified Ashworth scale. Due to limitations associated with patient access and cost, repeat scoring by independent evaluators was not possible. Plasma samples for the Control group were obtained from Mass General Brigham Biobank, a biorepository of consented patients’ samples at Mass General Brigham (parent organization of Massachusetts General Hospital and Brigham and Women’s Hospital). The sample cohort includes 38 individuals selected from the 20,270 available plasma samples of patients aged 0-25 **(Supplementary Table 1)**. The samples were selected based on four factors: (1) the age of the patient, (2) the Charlson Age-Comorbidity Index, (3) medication history, and (4) the history of medical problems. Samples from subjects with the Charlson Age-Comorbidity Index of 0-2 (corresponding to 98.3%-90.15% 10-year survival rate), taking no medication such as immune stimulants, immune suppressants, hormones/synthetics/modifiers, central nervous system and cardiovascular medications, blood products/modifiers/volume expanders and having no medical problems, i.e. no family history of cancer, heart disease and other chronic diseases have been selected for this study.

### Blood Sample Collection and Processing

Blood was collected in K2EDTA-coated Purple/Lavender top vacutainers (BD 368047). Plasma was isolated within 4 hours from collected blood samples by centrifugation at 1000g for 10 min at 4°C. Isolated plasma was stored at -80°C.

### SomaScan Proteomic Assay

Protein quantification was performed using SomaScan assay v.4.1 by SomaLogic (SomaLogic Operating Co., Inc., https://somalogic.com/). This is a high-throughput, multiplexed aptamer-based proteomic technology capable of measuring approximately 7,000 unique human protein analytes. Plasma samples from MLIV and control subjects were sent to SomaLogic Inc. on dry ice and analyzed in a single batch. Somalogic normalized the data for hybridization, intraplate median signal normalization, and plate scaling.

### Animal Studies and Sample Collection

*Mcoln1*^-/-^ mice were maintained as previously described (21). Genotyping was performed by Transnetyx using real-time qPCR (www.transnetyx.com). The *Mcoln1*^+/-^ breeders for this study were obtained by backcrossing onto a C57BL/6J background for more than 10 generations. Experimental cohorts were obtained from *Mcoln1*^+/-^ x *Mcoln1*^+/-^ mating. *Mcoln1*^+/+^ littermates were used as controls. Experiments were performed according to the Institutional and National Institutes of Health guidelines and approved by the Massachusetts General Hospital Institutional Animal Care and Use Committee.

### Statistical and Multivariate Analyses

#### Differential Expression Analysis

Differential expression analysis based on relative protein expression results from Somascan was performed with the limma package (v3.54.2) in R. This package was used to fit a linear model to the data and calculate empirical Bayes statistics. Prior to model fitting, variance stabilizing normalization was conducted with the normalizeVSN function, which log2 transformed the data. An FDR-adjusted p-value <0.05 was considered significant. Volcano plots were constructed using ggplot2 (v3.4.1) in R. Log2 fold-change thresholds were determined as the 15^th^ percentile of fold-change values for upregulated and downregulated with an FDR-adjusted p-value <0.05.

#### Gene Set Variation Analysis

To further elucidate functional changes, gene set variation analysis (GSVA) was performed (23). This analysis was conducted using the GSVA package (v1.44.5) available in R through Bioconductor with the relative protein expression values. Significant differences in gene set enrichment scores between the two groups were determined via gene set permutation analysis, as we have done before (24). Gene labels were randomly shuffled, and GSVA was recalculated. This procedure was repeated 1000 times, and p-values were computed as the rate that a random difference in gene set means between groups was greater in magnitude than the true difference. Benjamini and Hochberg’s false discovery rate adjusted p-values <0.05 were considered significant. The gene sets used for this analysis were the human Reactome Database and Pathway Interaction Database (PID) obtained from MSigDB C2 gene sets. A minimum gene set size of 10 was used, leading to a total gene set count of 1104.

#### Correlation Analysis

Plasma proteins and gene sets were correlated with motor scores to elucidate potential biomarkers of patient motor function. Spearman’s correlation was selected due to the expected nonlinear but monotonic response between protein expression and motor function. Correlation statistics were obtained using the “cor” function within the stats (v4.2.3) package in R. Relative expression values were used to correlate individual proteins, and enrichment scores from GSVA were used to correlate gene sets.

#### Gene Ontology Analysis

To provide specific functional annotations on correlated clusters of DEPs, gene ontology (GO) analysis was performed using PANTHER (v8) (25). Protein names were loaded using a csv file for a statistical overrepresentation test. The reference list was set as the 7322 proteins identified in this analysis. Significant terms were determined as those with an FDR-adjusted p-value <0.05 following a Fisher’s Exact test.

### Gene Functional Annotation

To provide additional functional annotation of the DEGs, a web-scraper was built to parse the National Center for Biotechnology Information Gene Database entry for each transcript and identify relevant terms. Transcript names were connected to the NCBI Entrez ID with the Genome wide annotation for Human package, org.Hs.eg.db (v3.15.0), available through Bioconductor in R This tool utilized the HTML parser in the BeautifulSoup4 (v4.12.2) Python package to extract the gene summary information from each gene entry in the database. The script then identified the keywords in the summary.

## Results

### Mucolipidosis Type IV Patients show Differential Blood Plasma Protein Signatures

We began the current study by asking if we could identify distinctive plasma protein signatures using the SomaScan assay (**Fig. 1A**). To do so, we analyzed protein content in plasma samples from 17 MLIV patients (**Table 1**) and 38 age-matched Biobank controls (**Supplementary Table 1**). The assay detected 7322 proteins and revealed one cluster of up-regulated proteins in MLIV patients and a second cluster that was down-regulated in MLIV patients (**Fig. 1B, Supplementary Table 2**). Differential analysis of protein abundances between MLIV and control groups revealed 1961 differentially expressed proteins (DEPs, **Fig. 1C**, adjusted p <0.05, and log_2_-foldchange >0.29 or log_2_-foldchange <−0.22, **Methods**, **Supplementary Table 3**). To gain insight into the abundance of DEPs associated with established functions relevant to MLIV, we built an NCBI database scraper to assign MLIV-relevant annotations to each of the DEPs (autophagy, interferon, iron, lipid, muscle, neuron, see **Methods**, **Supplementary Table 3**). Labeling these annotations on the volcano plot revealed that DEPs in all annotation groups were found among both upregulated and downregulated signatures (**Fig. 1C,D**). Importantly, we found a higher percentage of upregulated DEPs in “Interferon”, “Lysosome” and “Muscle” annotation groups, whereas more downregulated DEPs were found in “Neuron/Neural” annotation group. Given that these annotations were derived from our understanding of MLIV pathophysiology, these data suggest that the plasma contains protein signatures relevant to the tissue compartments and mechanisms involved in MLIV.

**Figure 1:**
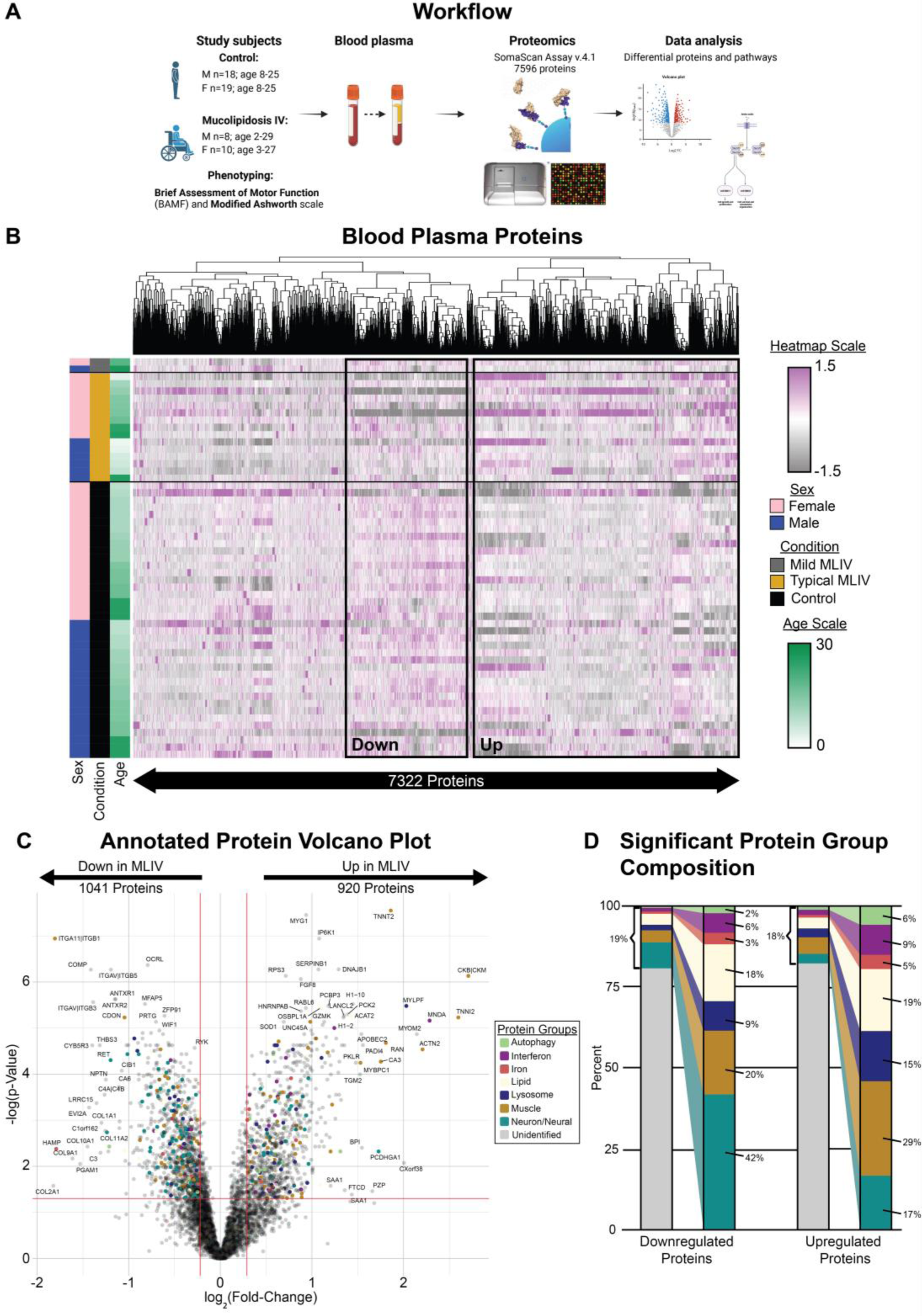
Proteomic profiling of blood plasma in MLIV and age-matched controls. **(A)** Hierarchical clustering of blood plasma proteins quantified in MLIV patients and age-matched controls (columns are z-scored). (**C**) Differential analysis revealed elevated proteins in MLIV or CTRL subjects (Wilcoxon rank sum). Proteins are color-annotated for MLIV-relevant functions based on NCBI-scraper. (**D**) MLIV-relevant annotations represent 18% of all up-regulated and 22% of all down-regulated protein.

**Table 1:** Clinical and genotype data for MLIV study subjects.

| Patient | Age (years) | Mode of Diagnosis | Phenotype | *MCOLN1* Genotype |  |
| --- | --- | --- | --- | --- | --- |
| 1 | <5 | Genetic | Typical | c.304 C>T | c.1405A>G |
| 2 | <5 | Genetic | Typical | c406 -2A>G | c.1336G>A |
| 3 | <5 | Genetic | Typical | c.1017_1020delACG<br>G | c.877 G>A |
| 4 | 5-10 | Genetic | Typical | c.984 +1G>A | c.406 -2A>G |
| 5 | 5-10 | Genetic | Typical | c.406 -2A>G | c.964C>T |
| 6 | 5-10 | Genetic | Typical | c.694A>C | c.785T>C |
| 7 | 11-15 | Genetic | Typical | c.406 -2A>G | c.406 -2A>G |
| 8 | 11-15 | Genetic | Typical | c.406 -2A>G | c.406 -2A>G |
| 9 | 11-15 | Genetic | Typical | c406 -2A>G | c.406 -2A>G |
| 10 | 11-15 | Genetic and Corneal bx | Typical | c406 -2A>G | Not Identified |
| 11 | 16-20 | Genetic | Typical | c.236_237ins93 | c.694A>C |
| 12 | 16-20 | Genetic | Typical | c406 -2A>G | c406 -2A>G |
| 13 | 16-20 | Genetic | Mild | c.236_237ins93 | c.1207C->T |
| 14 | 25-30 | Clinical and Gastrin | Typical | Unknown | Unknown |
| 15 | 25-30 | Genetic | Typical | c.406 -2A>G | c.406 -2A>G |
| 16 | 25-30 | Genetic | Typical | c406 -2A>G | c406 -2A>G |
| 17 | 25-30 | Genetic | Mild | c.920delT | c.1615delG |

### Plasma Proteins in MLIV Patients Correlate with Motor Function Scores

In MLIV patients, we simultaneously assessed motor function via the Brief Assessment of Motor Function scales (BAMF) and muscle tone via the modified Ashworth scale as previously described (2). Thus, we next asked if motor function or muscle tone is associated with specific blood protein signatures in MLIV patients. To test this, we conducted Spearman correlations between the 1,961 DEPs in MLIV patients (either up- or down-regulated) and key motor function (BAMF) and muscle tone (Ashworth) scores as well as age (**Fig. 2A,B** red indicates correlation with age or diminished function, **Supplementary Table 4**). Among the up-regulated DEPs, this analysis revealed a cluster of 212 proteins that “progressively” increased with worsening muscle hypertonicity, Gross Motor and UEGMS scores, and increasing age (**Fig. 2A, Cluster 1, Supplementary Table 5**). Gene ontology (GO) terms significantly over-represented in Cluster 1 included muscle protein development, TAK1 activation, and extracellular exosome. A second cluster of 254 proteins (**Cluster 2**) was associated with better muscle tone assessed via Ashworth and decreased with age, suggesting an adaptive or possibly “fatigued” up-regulation with disease progression. GO terms for Cluster 2 included mitochondrion, lysosomal membrane, protein translation, and RNA binding (**Fig. 2A**). Among the downregulated DEPs (**Fig 2B, Supplementary Table 5**), this analysis revealed a cluster (**Cluster 3**) of 588 proteins positively correlated with degree of hypertonicity and age, suggesting an early potentially adaptive downregulation that is lost with disease. GO terms enriched in Cluster 3 proteins included axon development, signaling, and cell adhesion. Among downregulated DEPs, a second cluster of 139 “progressive” down-regulated proteins was correlated with better muscle tone and motor function and decreased with patient age (**Cluster 4**). GO analysis of proteins in this cluster identified functions associated with neuronal development and extracellular matrix (ECM). These findings reveal proteomic signatures consistent with our understanding of MLIV’s natural history, including worsened neuronal function and increasing involvement of metabolic proteins with disease progression.

**Figure 2:**
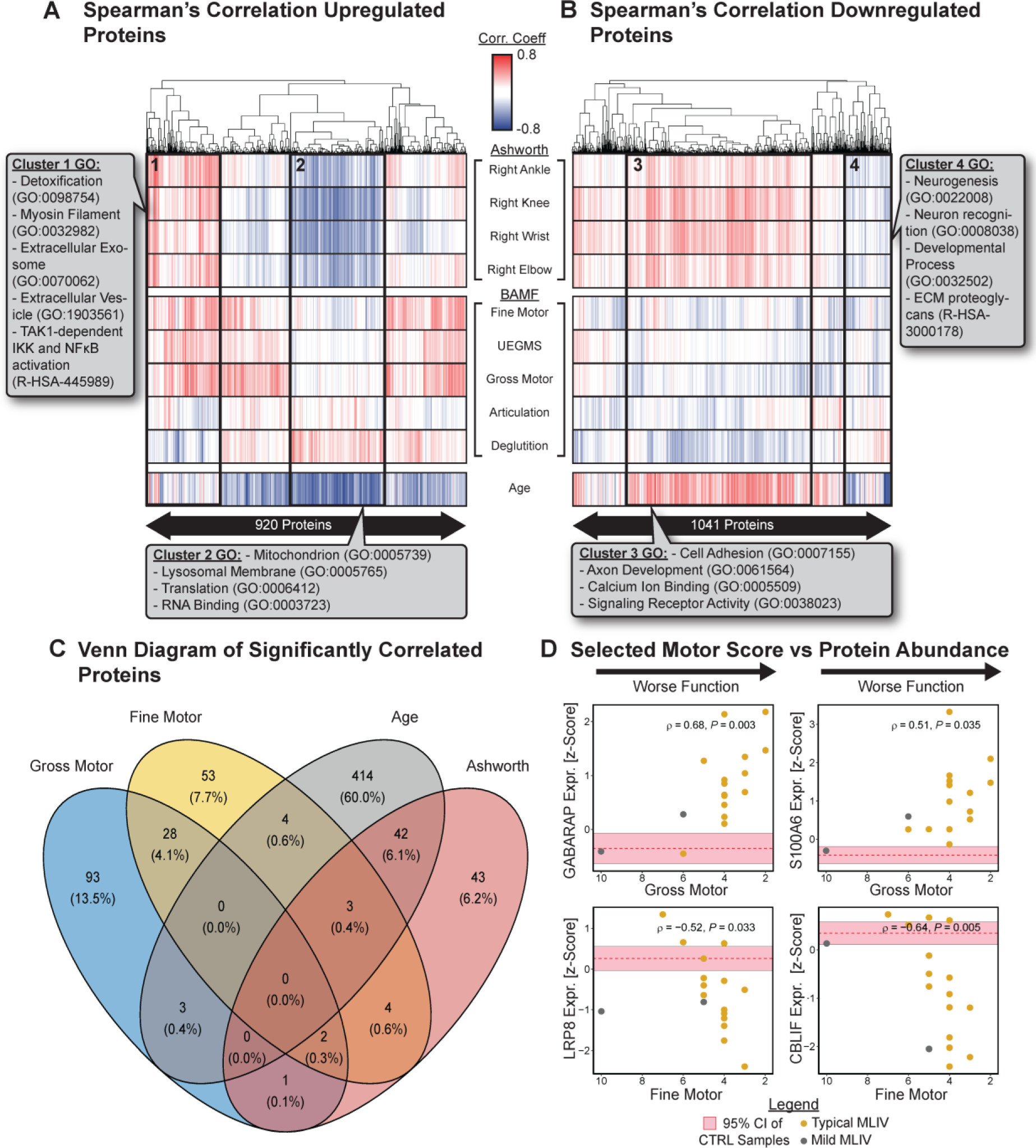
Correlation of differentially expressed proteins with clinical scores. (**A**) Spearman correlation coefficient between MLIV up-regulated DEPs and clinical scores (hierarchical clustering). (**B**) Spearman correlation coefficient between MLIV down-regulated DEPs and clinical scores (hierarchical clustering). (**C**) Venn diagram of a number of significantly correlated proteins with selected clinical scores (gross motor, fine motor, at least two Ashworth measures) and age. (**D**) Correlation of selected significantly correlating proteins with clinical scores in MLIV patients (proteins z-scored, Spearman correlation computed across all MLIV samples, un-adjusted p-value). Orange dots are typical MLIV cases; gray dots are mild MLIV cases.

To better understand how the individual DEPs of MLIV patients correlate with different functional variables, such as gross motor and fine motor function, muscle tone (Ashworth), and age, and whether some of the proteins can overlap between these functional variables, we created a Venn diagram (**Fig. 2C, Supplementary Table 6**). Proteins were considered to correlate with Ashworth scores if there was significant correlation with at least two individual scores. Importantly, we found that the majority of the significant correlations were with age, indicating age-related “progressive” protein signature of the disease (2, 26). Among all DEPs, only GABA Type A Receptor-Associated Protein (GABARAP) and S100 Calcium Binding Protein A6 (S100A6) were significantly correlated with gross motor, fine motor, and Ashworth scores (**Figs 2C,D**). GABARAP is a mediator of neurotransmitter receptors that correlates with age and disease severity (**Fig. 2D**). S100A6 is a calcium binding ion channel expressed by pyramidal neurons (27), and its increase in MLIV is correlated with disease severity. Additionally, LDL receptor related protein 8 (LRP8) was significantly correlated with both age and improved fine motor scores, in a negative and positive manner, respectively. This protein mediates neuroplasticity in the adult brain through interaction with Reelin (28). Similarly, cobalamin binding intrinsic factor (CBLIF), decreased with age and increased with fine motor function. This protein is critical for the sufficient intestinal absorption of vitamin B_12_ in the intestine (29), which is required for myelination and deficiency leads to neurologic pathology (30). All other proteins presented in the Venn diagram (**Fig. 2C**) are detailed in **Supplementary Table 6**.

### Gene Set Analysis Identifies Differentially Enriched Gene Sets in MLIV Plasma Associated with Brain Hallmarks of the Disease

Having identified up- and down-regulated DEPs, some of which correlated with either clinical scores or age (**Figs 1,2**), we next asked if we could identify signatures from existing gene set annotations using the same blood proteomic profiling. To test this, we used gene set variation analysis (GSVA) (23) together with gene sets from the Reactome and Pathway Interaction Databases (PID), sourced from MSigDB. Doing so revealed 229 significantly enriched gene sets in MLIV samples (upregulated in MLIV) and 22 significantly enriched gene sets in Controls (downregulated in MLIV) (**Fig. 3A**). The gene sets enriched in MLIV, included sphingolipid biosynthesis, MAPK and AKT signaling, interleukin 1, and autophagy (**Fig. 3A**, **Supplementary Table 7**), all of which are consistent with our prior findings from pathological and proteomic analysis of brains in the MLIV mouse model (31–34). Because GSVA collapses genes into a gene set enrichment score for each subject, we next asked if we could identify pathways that are correlated with muscle tone and motor function scores, similarly to our analysis of proteins in **Fig. 2**. This revealed a cluster of significantly enriched gene sets that correlated with poorer muscle tone and motor function (**Fig. 3B, “Ashworth – Up” cluster, Supplementary Table 8, 9**)– these included gene sets involved in growth factor and MAPK pathway signaling, synaptic signaling, and autophagy. A second cluster of gene sets correlated with better muscle tone and decreased with age (**Fig. 3B, “Ashworth – Down” Cluster**) and included gene sets associated with proteinase function, RUNX signaling, and MAPK signaling (**Supplementary Table 9**). Among these correlations, 14 gene sets were significantly correlated with at least two gross motor, fine motor, and Ashworth scores (**Fig 3C, Supplementary Table 10**). All 14 of these gene sets were positively correlated with clinical scores (**Fig. 3D**), indicating their enrichment scores decrease with disease severity. Their functions span immune signaling, mitosis, and cell cycle, reflecting an overall dysregulation of cellular function with clinical severity.

**Figure 3:**
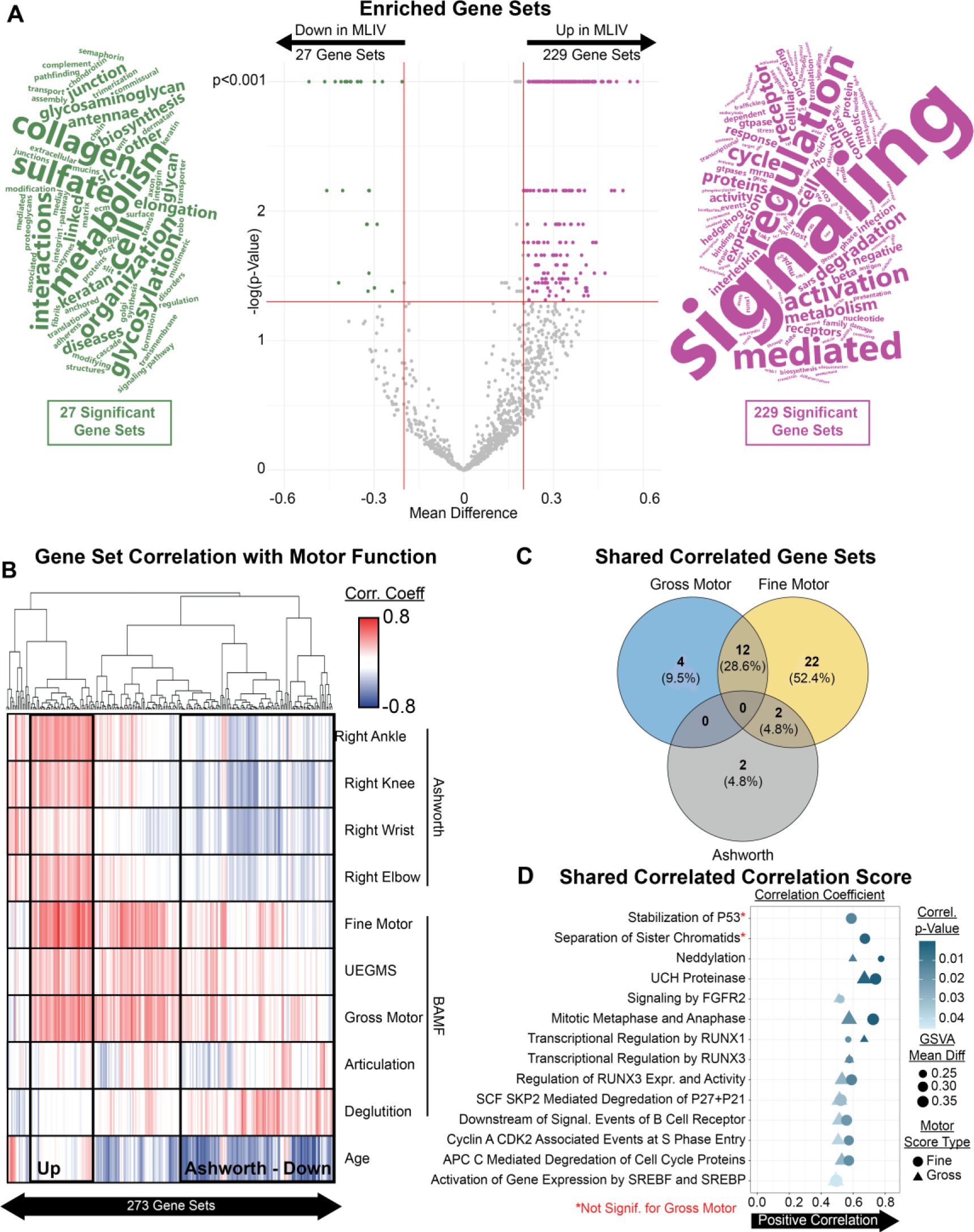
Gene Set Analysis Reveals Enrichment of MLIV Signatures in Plasma. (**A**) Gene set variation analysis using Reactome and PID gene set libraries identifies 299 gene sets enriched in MLIV samples and 27 gene sets enriched in Control samples (pFDR<0.05). Adjacent word clouds show relative frequency of terms in gene set names from the significant regions of volcano plot. (**B**) Correlation of gene set scores with motor function, muscle tone, and age reveals gene sets correlated with worse functional scores (“Up” cluster) or correlated with better muscle tone and decreasing with age (“Down” cluster). (**C**) Among the gene sets that reach significance (p<0.05), 14 significantly correlated with at least two of fine motor, gross motor, and Ashworth scores. (**D**) Enrichment of 14 gene sets significantly positively correlated with fine motor and gross motor scores.

### Human Plasma Proteome Shares DEPs with MLIV mouse brain tissue

Because MLIV patient brain samples are not available, our research group recently used mass spectrometry to profile the brain proteome in the mouse model of MLIV (32, 35). Armed with these data (**Supplementary Table 11**), we next asked if we could identify shared signatures between the mouse MLIV brain and human plasma. Among the 197 brain DEPs and 1961 plasma DEPs, we found that 45 overlapped between datasets (**Fig. 4A**,**B, Supplementary Table 12**). The up-regulated overlapping proteins were largely related to lysosomal function, while the down-regulated proteins were largely related to myelination. Both of these signatures are consistent with our understanding of key disease hallmarks: impaired myelination and modified lysosomal activity. Importantly, GSVA of both datasets using the same Reactome and PID gene sets used above (**Fig. 2**) also revealed shared up-regulated immune pathways in MLIV human plasma and MLIV mouse brain (**Fig. 4C,D**, **Supplementary Table 13**), suggesting immune changes in both compartments. Collectively, these data emphasize the probable presence of shared features between the brain and the periphery. Because an abundance of proteins are shared between regions and move in the same direction, and many of them are related to the primary pathology found in the MLIV brain, e.g, down-regulation myelination-related proteins and up-regulation of lysosomal proteins; our findings support the presence of a plasma protein profile that reports brain pathology. Thus, this work provides a panel of plasma proteins that may be used in future studies for the validation of plasma biomarkers of MLIV.

**Figure 4:**
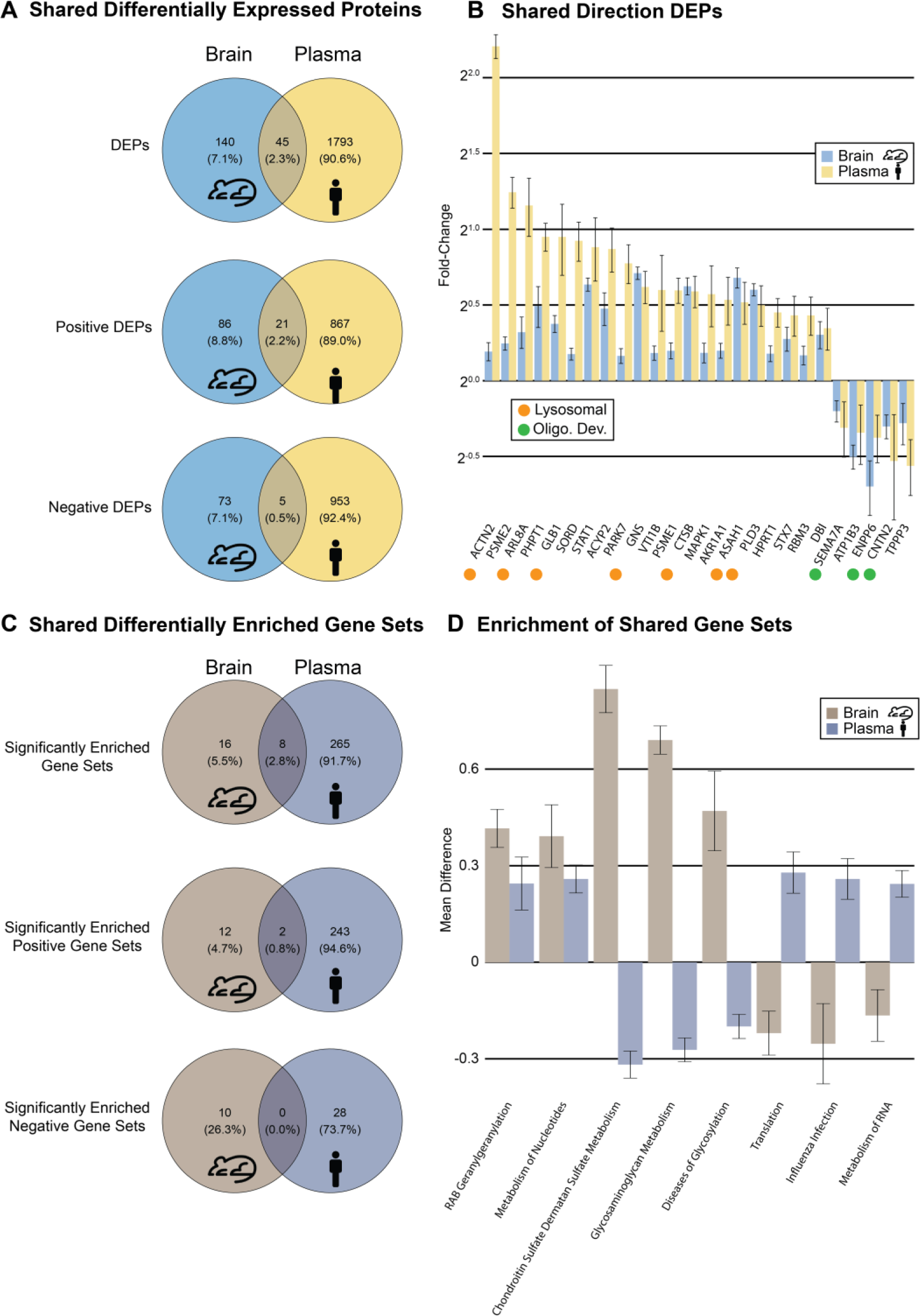
Human plasma and mouse brain share MLIV protein signatures. (**A**) Shared and distinct DEPs in human MLIV plasma and mouse MLIV brain. (**B**) DEPs with the same direction in human plasma and mouse brain (pFDR<0.05). Annotations indicate the presence of literature connecting that protein with either lysosome function or oligodendrocyte development. Fold change greater than 2^0^ indicates higher expression in MLIV patients/model system relative to control. (**C**) Shared and distinct significantly enriched gene sets by in human plasma and mouse brain. (**D**) Shared significantly enriched gene sets in human plasma and mouse brain (permutation pFDR<0.05). Positive mean difference indicates greater enrichment in MLIV patients/model system relative to control.

## Discussion

In this study, we comprehensively profiled proteomic differences in blood plasma between MLIV patients and bio-banked controls and correlated differential proteins with clinical motor function and muscle tone scores.

Motivated by our prior work showing shared cytokine changes in MLIV between human blood and mouse brain (26), we hypothesized in the current study that that we would identify blood signatures associated with the primary neuronal and lysosomal dysregulation found in the MLIV brains. Because data is limited in human MLIV brains, we compared our findings in human blood with mouse MLIV brain proteome, identifying a subset of shared changes between these compartments/species. Importantly, many of the overlapping proteins were involved in lysosomal function (**Fig. 4**). This study is the first to define changes in the blood proteome and identify those that are consistent with the MLIV brain. It thus represents an essential first step in the identification of clinically relevant blood biomarkers of MLIV.

Recently, we reported an AAV-MCOLN1 gene replacement strategy that restored motor function when administered to presymptomatic and symptomatic *Mcoln1^-/-^* mice. Excitingly, these data suggest that *MCOLN1* gene therapy may be able to restore motor development in patients rather than simply delaying disease progression. However, the time needed for restored developmental processes to produce a clinically meaningful improvement in function in humans is uncertain and may exceed the trial timeframe. As such, there is a critical unmet need for a clinically tractable biomarker to measure TRPML1 activity restoration in the brain. MLIV human brain tissue analysis has been historically restricted by the lack of tissue donation and is currently limited to only three published cases (7, 8, 33).

Our findings of multiple lysosomal plasma proteins associated with clinical severity or overlapping between brain and plasma are important because they may directly reflect the central role of MCOLN1 in lysosomal regulation. The protein encoded by MCOLN1 is a lysosomal non-selective cationic channel from the transient receptor potential family, TRPML1, or mucolipin-1. TRPML1 is a versatile regulator of lysosomal function. Many of these functions are attributed to its Ca2+-transport from lysosomes, including Ca^2+^-dependent lysosomal membrane fusion events in endocytosis, exocytosis, and autophagy (14); regulation of the transcription factor EB and the CLEAR gene network (15), autophagosome membrane assembly (16), and chaperon-mediated autophagy (17). More recently, TRPML1 was also shown to regulate Conjugation of ATG8 to Single Membranes, or CASM, a non-canonical form of autophagy induced by lysosomal damage, LC3-associated endocytosis or phagocytosis (36). In addition to these experimentally established functions of TRPML1 facilitated via transport of Ca2+, its role in lysosomal transport and maintaining homeostasis of iron and zinc has also been documented (37–39).

To identify protein signatures associated with clinical manifestations of MLIV, we have performed a correlation analysis of BAMF (gross motor function, fine motor function, upper extremities gross motor function, articulation, and deglutition), modified Ashworth scores for muscle tone collected in patients and their age. We found a significant correlation of up and downregulated proteins with each of these functional variables, as indicated in **Fig. 2**. Notably, we have identified clusters of up- and downregulated proteins associated with clinical measures attributed to disease severity, shown as Clusters 1-4 (**Fig. 2**), and GO analysis identified GO terms and processes enriched in these clusters.

Proteins in Cluster 1 represent the group that is upregulated in MLIV as compared to healthy controls and positively correlate with disease severity, i.e., worsening of the fine and gross motor function and muscle rigidity. GO analysis of this protein signature showed enrichment in metabolic process, detoxification, extracellular vesicles, and toll-like receptor signaling. Proteins in Cluster 2 are on average upregulated in the plasma of MLIV patients compared to healthy controls and decrease with worsening muscle tone and age. In contrast, Cluster 4 shows the group that, on average, is downregulated in MLIV but increases with age and worsening muscle tone. Thus, both Clusters 2 and 4 may demonstrate a “fatigued” response with age. It is interesting to note that protein patterns in Clusters 2 and 4 showed that many of the proteins that changed in MLIV plasma with age were associated with increasing rigidity (i.e worsening muscle tone). This pattern was absent or less pronounced for BAMF measures (i.e gross/motor function, articulation and deglutition). These observations are supported by age-correlation data of the neurological assessments in a cross-sectional natural history study (2), where muscle tone rather than BAMF measures showed a significant correlation with patients’ age. The signature in Cluster 2 (upregulated and declining with age and rigidity) showed enrichment in mitochondrion, lysosomal membrane, RNA binding and regulation of translation. In contrast, in Cluster 4 (downregulated but increasing with age and rigidity), it was enriched in neurogenesis, axon development, Ca2+-binding and cell adhesion. Proteins in Cluster 3 are, on average, downregulated in MLIV and declined with worsening muscle tone and fine motor function. GO terms enriched in this group were related to nervous system development, synaptic membrane, and lysosomal lumen.

Interestingly, we found several significantly correlated proteins that overlapped between different clinical measures. For example, higher abundancies of GABARAP and S100A6 were commonly correlated with worse clinical presentation in gross motor function, fine motor function, and muscle tone (**Figs. 2C,D**). GABARAP, together with GABARAPL1, 2, and 3, form a family of GABA receptor-associated proteins that, with LC3 proteins, are a part of the ATG8 family, conjugation proteins that are essential for autophagy. Together with the well-established and overlapping functions between LC3 and GABARAPs in autophagophore formation, GABARAPs have a broad array of unique functions, both in autophagy and autophagy-unrelated that have been recently extensively reviewed (40–43). Autophagy-related functions of GABARAPs, distinct from LC3, include autophagy initiation, cargo targeting, autophagosome-lysosomal fusion, TFEB-dependent coordination of lysosomal biogenesis and capacity and selective autophagy, such as mitophagy, ERphagy, as well as autophagic degradation of Golgi and centrosomes. Outside of autophagy, GABARAPs, as implied by their name, are involved in the binding and transport of GABA receptors and, as such, regulation of synaptic transduction and nerve signaling (44). Interestingly, in a recent study, GABARAPs were shown to be essential for TRPML1-mediated activation of TFEB, providing new evidence of mechanistic relationships between the two proteins (36).

GABARAPs were reported to be altered in several human diseases. Expression of GABARAPs (particularly GABARAP and L1 forms) is inhibited in different forms of cancer, and they are hypothesized to act as tumor suppression factors (reviewed in (40)). In multiple myeloma, breast cancer and some other forms, GABARAP expression is used for risk, detection, and prognosis, and treatment efficacy evaluation. Changes in GABARAPs expression have also been associated with neurodegenerative diseases, including Stiff person syndrome (45), Dementia with Lewy bodies (46), (47), epilepsy (48), and neurodevelopmental syndrome due to chromosomal 17p13.1 microduplication (49).

Our data show that all members of the GABARAP family were significantly upregulated in MLIV plasma (**Supplementary Table 3**), and, importantly, two members, GABARAP and GABARAPL1, positively correlated with worsening gross motor, fine motor, and muscle tone (the former) or gross motor and fine motor (the latter) clinical scores in MLIV patients. It is currently unknown whether the increase in GABARAPs in MLIV is due to global inhibition of autophagy or whether it reflects CNS-driven dysregulation of GABA receptor trafficking. The elucidation of the detailed mechanism was out of the scope of the current study. Yet, the consistent increase of GABARAPs and their significant correlation with disease severity make them strong candidates for follow-up validation as biomarkers.

Another protein that is significantly associated with the worsening of gross and fine motor function and muscle tone identified in the plasma of MLIV patients in our study is S100A6 or calcyclin. It is a Ca^2+^ and Zn^2+^-binding secreted protein with many functions related to cell proliferation and differentiation, cytoskeleton reorganization, and stress response (50). Changes in secreted S100A6 in body fluids were found to be associated with diseases. Increased serum S100A6 positively correlates with the progression of different forms of cancer and is suggested to be used as a biomarker. The increase of S100A6 is also linked to fibrosis-related conditions such as liver cirrhosis biliary, chronic renal disease, pulmonary fibrosis, and myocardial infarction. Additionally, high S100A6 is also linked to astrogliosis in AD and ALS (27, 51). The direct mechanistic relationships between loss of TRPML1 and upregulation of S100A6 have not been established, but due to association with the broad spectrum of clinical manifestations in MLIV and its established mechanistic links to Ca2+/Zn2+ regulation and astrocytic pathology in AD and ALS, we conclude that S100A6 can be further evaluated as disease severity markers of MLIV.

We also found that three proteins, LRP8 or ApoER2, CBLIF, and SHMT1, were commonly associated with worse muscle tone, fine motor function, and age in MLIV plasma and, therefore, can also be evaluated as potential disease severity markers of MLIV. Of these three, ApoER2 and CBLIF showed a negative correlation with disease severity, and SHMT1 positive. ApoER2 is one of two receptors that bind reelin and facilitate down-stream signaling known for its role in neuron migration in brain development and synaptic modulation (52). Progressive reduction of CBLIF, a cobalamin-binding intrinsic factor, which is required for cobalamin or vitamin B12 absorption in the ileum, warrants evaluation of MLIV patients for cobalamin deficiency and, if needed, supplementation. SHMT1 is a cytosolic serine hydroxymethyltransferase, that has been linked to cancers and cardiometabolic disorders with aging (53, 54).

To reveal a plasma signature of MLIV related to pathologic alternations in the brain, we have compared protein signatures of DEPs in the plasma of MLIV patients and MLIV mouse brains. Specifically, this comparative analysis with the whole brain tissue (32, 35) showed concordant protein expression through a downregulation of proteins related to oligodendrocyte development and myelination, and an upregulation of lysosomal proteins. Our data identified 21 upregulated and 5 downregulated DEPs, overlapping between the MLIV patient’s plasma and MLIV mouse brain tissue. These proteins present a set of biomarker candidates for MLIV for future validation. Among upregulated, 7 (ACTN2, ARL8A, GLB1, GNS, CTSB, ASAH1, and PLD3) were lysosomal proteins; out of 5 overlapping downregulated DEPs, 3 (SEMA7A, ENPP6, and CNTN2) were related to oligodendroglia. This pattern highlights the similarity between mouse brain and human plasma protein signatures in MLIV. Other upregulated DEPs that overlapped in MLIV human plasma and mouse brain were mitochondrial proteins (ACYP2, SORD, PARK7 and MPST) or involved in proteasomal protein degradation (PSME2, PSME1). Species. This study is the first to define changes in the blood proteome and identify those that are consistent with the MLIV brain model. It thus represents an essential first step in the identification of clinically relevant blood biomarkers of MLIV.

We selected the SomaScan platform because of its reported broad dynamic range providing the capacity to analyze blood plasma (55). Indeed, we were able to identify brain-related signatures in the blood, including myelin, neurons, etc, as well as proteins associated with lysosomal function. We emphasize that this is an early discovery study, representing a screen of more than 7,000 proteins across a cohort of just 17 MLIV cases. Nevertheless, our findings here, including more than 800 DEPs with a positive correlation with age and 26 proteins with overlapping changes between human blood and mouse brain, represent a strong foundation to inform future biomarker validation studies. We are strongly encouraged by our observation that many of the DEPs in blood plasma are brain proteins and are relevant to MLIV pathogenesis, e.g., myelin, etc. One important limitation is that the clinical scores and blood samples were taken as a single snapshot of each patient at different stages rather than a longitudinal analysis. Moreover, the clinical scores are limited in assessing the overall level of neurological dysfunction (2). Nevertheless, natural history data is broadly limited for MLIV, and this work represents the first study to systematically connect clinical phenotypes with underlying molecular fingerprints of the disease.

In conclusion, this study defines the first blood proteomic profile of MLIV and the first analysis connecting blood proteins to both clinical scores and a brain proteomic signature, albeit in the mouse. Our analysis identified one subset of markers that are associated with disease severity and a second subset shown to be dysregulated in the brain. As discussed above, many of the proteins identified are directly implicated in neuronal and lysosomal function (e.g., GABARAP) as well as myelination, which are several of the central affected functions in MLIV. We posit that proteins associated with lysosomal dysfunction represent a highly promising biomarker panel for MLIV because these are directly connected to the molecular mechanism of loss of TRPML1 function in the disease. Future targeted studies should evaluate how these blood signatures change longitudinally and whether they are affected by forthcoming therapeutic studies.

## Author Contributions

Y.G. and L.B.W designed and oversaw the execution of the study. A.M. conducted motor function and clinical assessment of motor function scores. V.M-B. and M.S. coordinated human plasma and mouse brain sample and data collection; B.T. executed computational data analysis. B.T, Y.G., and L.B.W drafted the manuscript. All authors reviewed the manuscript.

## Data Availability

All data produced in the present work are contained in an anonymized form in the manuscript.

## Acknowledgments

We thank Mass General Brigham Biobank for providing control samples and health information data; the ML4 foundation, Dr. Rebecca Oberman and Randy Gold for providing research funding (Y.G) and help with MLIV blood samples collection. This work was also funded by the UPenn Orphan Disease Million Dollar Bike Run grant (to A.M. 2017D007934) and Georgia W. Woodruff School of Mechanical Engineering Faculty Fellowship at Georgia Tech (L.B.W.). B.T an L.B.W. were funded in part by the National Institutes of Health under grant number 1R01AG075820.

## Disclosures

Y. G. received research funding from the ML4 Foundation; A.M. received funding from UPenn Orphan Disease Million Dollar Bike Run Grant Program. L.B. Wood and other co-authors report no disclosures relevant to the manuscript.

## Notes

### Author Declarations

Institutional Review Board of Massachusetts General Hospital gave ethical approval for this work.

